# Changes in emergency department utilization in vulnerable populations after COVID-19 shelter in place orders

**DOI:** 10.1101/2023.10.25.23297561

**Authors:** Philip R. Wang, Akhil Anand, James Bena, Shannon Morrison, Jeremy Weleff

**Author notes:** Corresponding author: Jeremy Weleff, DO, Yale University School of Medicine, Department of Psychiatry 300 George St, New Haven, CT, 06511. **Funding sources/disclosures**: This research did not receive any specific grant from funding agencies in the public, commercial, or not-for-profit sectors. All authors have no conflicts of interest, financial or otherwise, to declare related to this manuscript. **Credit author statement:** Concept and design: PW, AA, JW. Acquisition of the data: JB, SM, AA, JW. Analysis and interpretation of the data: PW, AA, JB, SM, JW. Drafting of the manuscript: PW. Critical revision of the manuscript for important intellectual content: AA, JW, JB, SM. Statistical expertise: JB, SM, JW. Acquisition of funding: None.

## Abstract

**Purpose:** To compare emergency department (ED) utilization and admission rates for patients with a history of mental health (MH), substance use disorder (SUD) and social determinants of health (SDOH) before and after implementing COVID-19’s shelter-in-place (SIP) orders.

**Methods:** This was a retrospective, multicenter study leveraging electronic medical record data from 20 EDs across a large Midwest integrated healthcare system from 5/2/2019 to 12/31/2019 (pre-SIP) and from 5/2/2020 to 12/31/2020 (post-SIP). Diagnoses were documented in the patient’s medical records. Poisson and logistic regression models were used to evaluate ED utilization and admission rate changes.

**Results:** 871,020 total ED encounters from 487,028 unique patients were captured. 2,572 (0.53%) patients had a documented Z code for SDOH. Patients with previously diagnosed MH or SUDs were more likely to seek ED care after the SIP orders were implemented (RR: 1.20, 95% CI: 1.18 – 1.22, p<0.001), as were patients with SDOH (RR: 2.37, 95% CI: 2.19 – 2.55, p<0.001). Patients with both previously diagnosed MH or SUD and a documented SDOH had even higher ED utilization (RR: 3.31, 95% CI: 2.83 – 3.88, p<0.001) than those with either condition alone. Patients with MH and SUDs (OR: 0.89, 95% CI: 0.86 – 0.92, p<0.001) or SDOH (OR: 0.67, 95% CI: 0.54, 0.83, p<0.001) were less likely to be admitted post-SIP orders while patients with a history of diseases of physiologic systems were more likely to be admitted.

**Conclusions:** Vulnerable populations with a history of MH, SUD, and SDOH experienced increased ED utilization but a lower rate of hospital admissions after the implementation of SIP orders. The findings highlight the importance of addressing these needs to mitigate the impact of public health crises on these populations.

## Background

Patients with mental health (MH) and substance use disorders (SUD) face significant challenges in accessing healthcare equitably.^1^ In addition, social determinants of health (SDOH) profoundly impact healthcare utilization, delivery, quality of life, and mortality. Identifying these patients for targeted interventions is crucial^2^; however, inconsistent and non-standardized documentation of SDOH across healthcare systems hinders public health surveillance efforts of this population. To address this issue, the Centers for Medicare and Medicaid Services and other groups have proposed using Z-Codes to standardize the documentation of social needs, but the utilization of these codes remains limited.^3,4^

The COVID-19 pandemic has exacerbated health disparities and disproportionately affected individuals with MH, SUD, and SDOH.^5^ While research has shown that shelter-in-place (SIP) orders have led to increased ED visits for mental health and substance use reasons^6–8^, few studies have examined the impact on ED utilization and admissions in individuals with a prior history of MH or SUD. Furthermore, the effects of the COVID-19 pandemic on ED utilization and admissions by patients with SDOH have had been poorly studied thus far. The primary aim of this study is to investigate the changes in ED utilization and hospital admission rates among vulnerable populations with a history of MH, SUD, and SDOH before and after SIP orders across a large integrated healthcare system. A secondary aim was to characterize the frequency of utilization of Z-codes to identify patients with SDOH in the ED.

## Methods

### Patient population

Patients who visited any of 20 EDs ranging from large hospital-associated EDs to freestanding EDs across a large Midwest integrated healthcare system Ohio from 5/2/2019-12/31/2019 (designated as the “pre-SIP order” time period) and 5/2/2020-12/31/2020 (the “post-SIP order” time period) were identified. Demographics and diagnoses were extracted from the electronic medical record (EMR). MH and SUDs were identified using the International Classification of Diseases 10th Revision (ICD-10) codes; similarly, SDOH were identified in patients’ medical histories using respective Z-Codes in the ICD-10. Historical use of theses codes means that these diagnoses and SDOH were documented in the patient’s chart at any time, and not solely reliant on ED encounter documentation. This study was approved by Cleveland Clinic IRB.

### Statistical analysis

Poisson regression models were used to calculate absolute risk ratios (RR) to compare the total number of cases between the pre-and post-SIP periods. Logistic regression models were used to evaluate changes in admission rates between the two periods, and odds ratios (OR) were calculated.

## Results

### Patient demographics

871,020 total ED encounters from 487,028 unique patients were captured; 473,449 visits (54%) occurred in 2019, and 397,571 (46%) were in 2020. Our cohort was mostly White (65.4%), female (53.8%), privately insured (68.9%), and had a mean age of 46.0 ± 24.0. 2,572 (0.53%) patients had a documented Z code for SDOH. The most coded SDOH in ED encounters was problems related to housing and economic circumstances, followed by other problems related to the primary support group and other psychosocial circumstances (Supplemental Table 1). Compared to patients without SDOH, patients with a coded SDOH were more likely to be Black, younger, identify as male, and were more likely to be on government or self-pay insurance (Supplemental Table 2).

### ED utilization rates before and after shelter-in-place orders

Compared to before the implementation of SIP orders, patients with previously diagnosed MH or SUDs were more likely to seek ED care after the SIP orders were put in place (RR: 1.20, 95% CI: 1.18 – 1.22, p<0.001) (Table 1). Among patients with SDOH, those with a history of problems related to the primary support group (RR: 2.89, 95% CI: 2.54 – 3.29) or housing and economic circumstances (RR: 2.50, 95% CI: 2.23 – 2.80) were more likely to present to the ED after SIP orders. Patients with both previously diagnosed MH or SUD and a documented SDOH (RR: 3.31, 95% CI: 2.83 – 3.88, p<0.001) had even higher ED utilization than those with either condition alone.

**Table 1.**
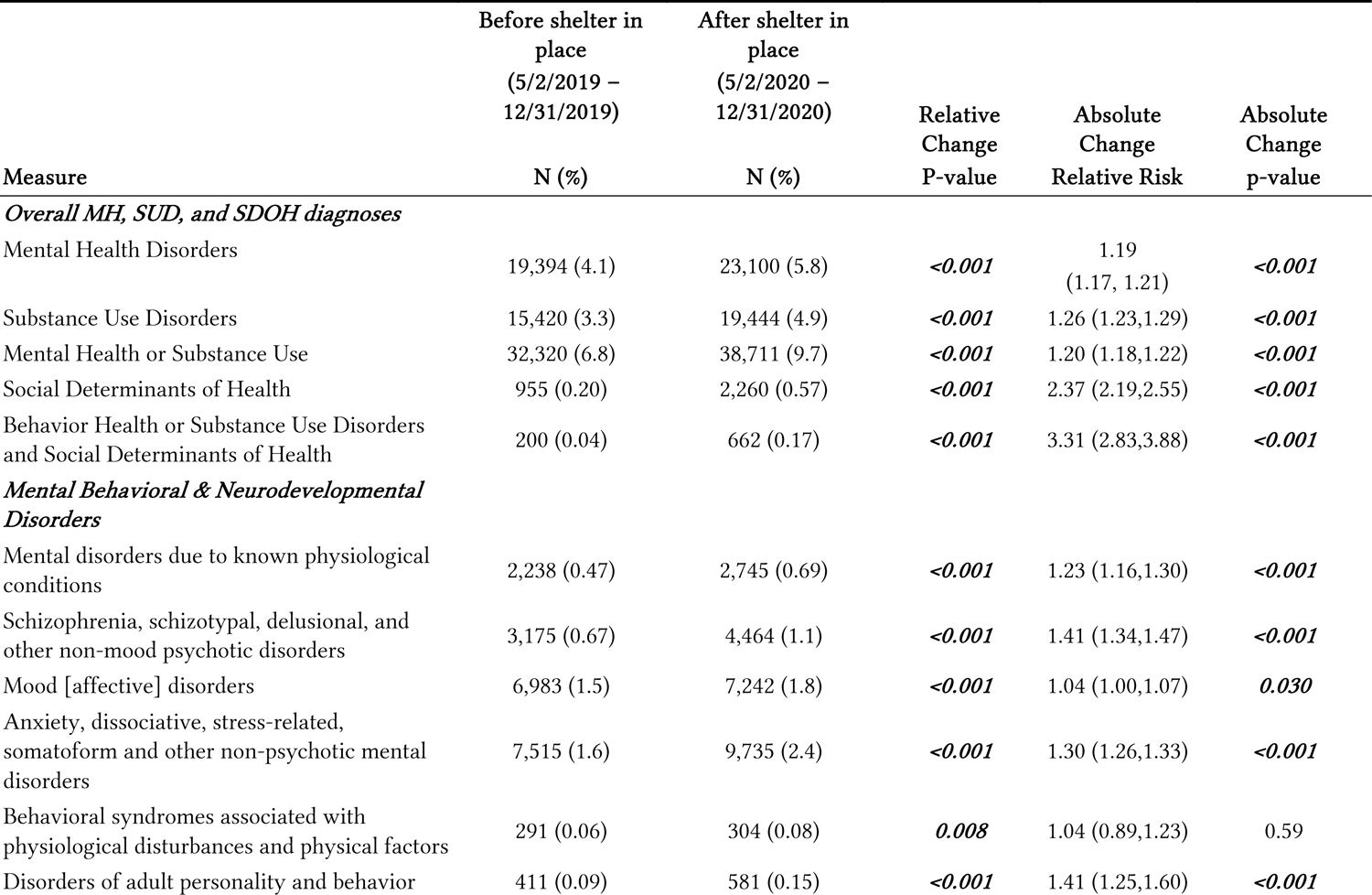
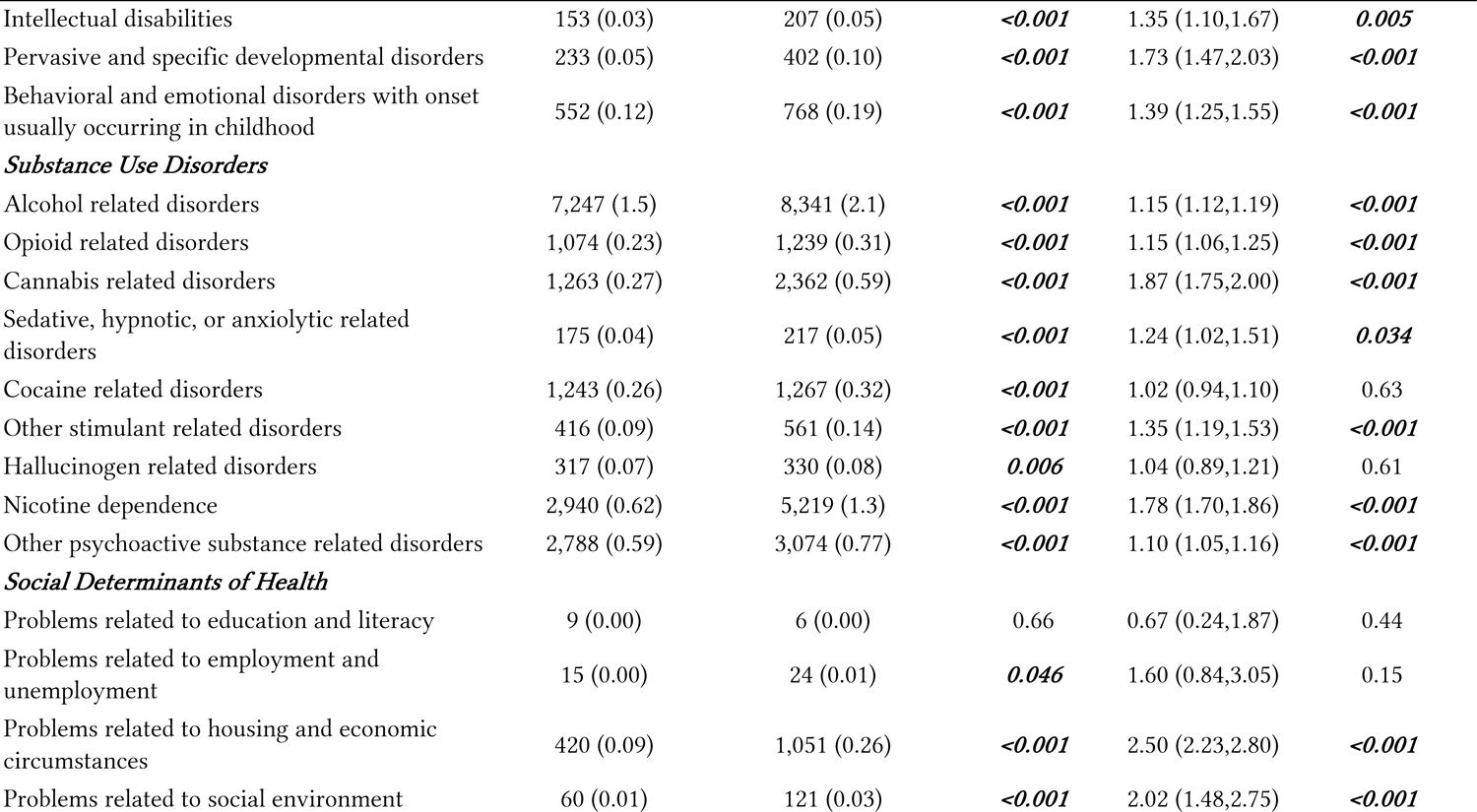
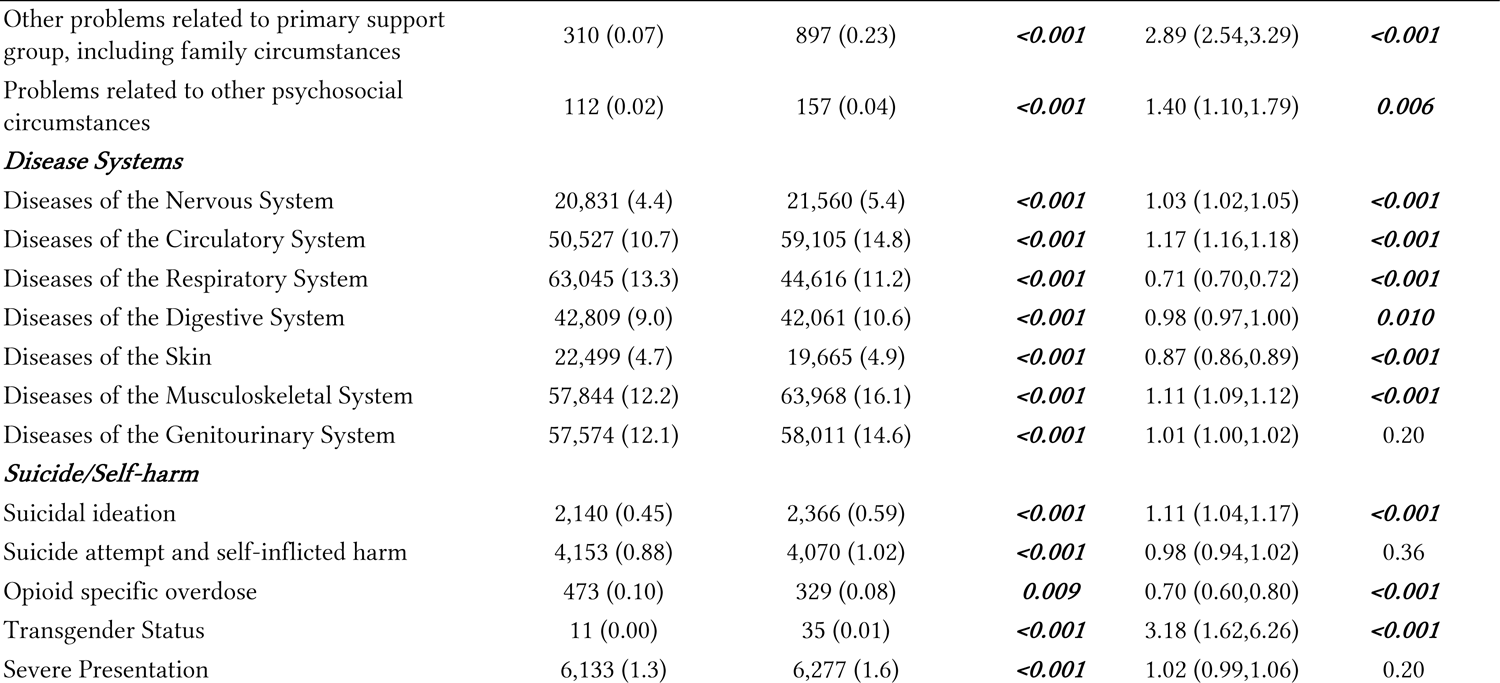
ED visits of categories of historical diagnoses before and after shelter-in-place orders.

### ED admission rates before and after shelter-in-place orders

Compared to pre-SIP, patients with a previously diagnosed MH or SUD (OR: 0.89, 95% CI: 0.86 – 0.92, p<0.001) or any documented SDOH (OR: 0.67, 95% CI: 0.54, 0.83, p<0.001) were less likely to be admitted to the hospital after presenting to the ED post-SIP orders (Table 2). Of the SDOH diagnosis categories, only patients with problems related to their primary support group and those with problems related to other psychosocial circumstances were significantly less likely to be admitted. The difference in admission rate pre- and post-SIP orders in patients with both MH/SUD and SDOH did not reach significance (OR: 0.65, 95% CI: 0.42, 1.02, p=0.061).

**Table 2.**
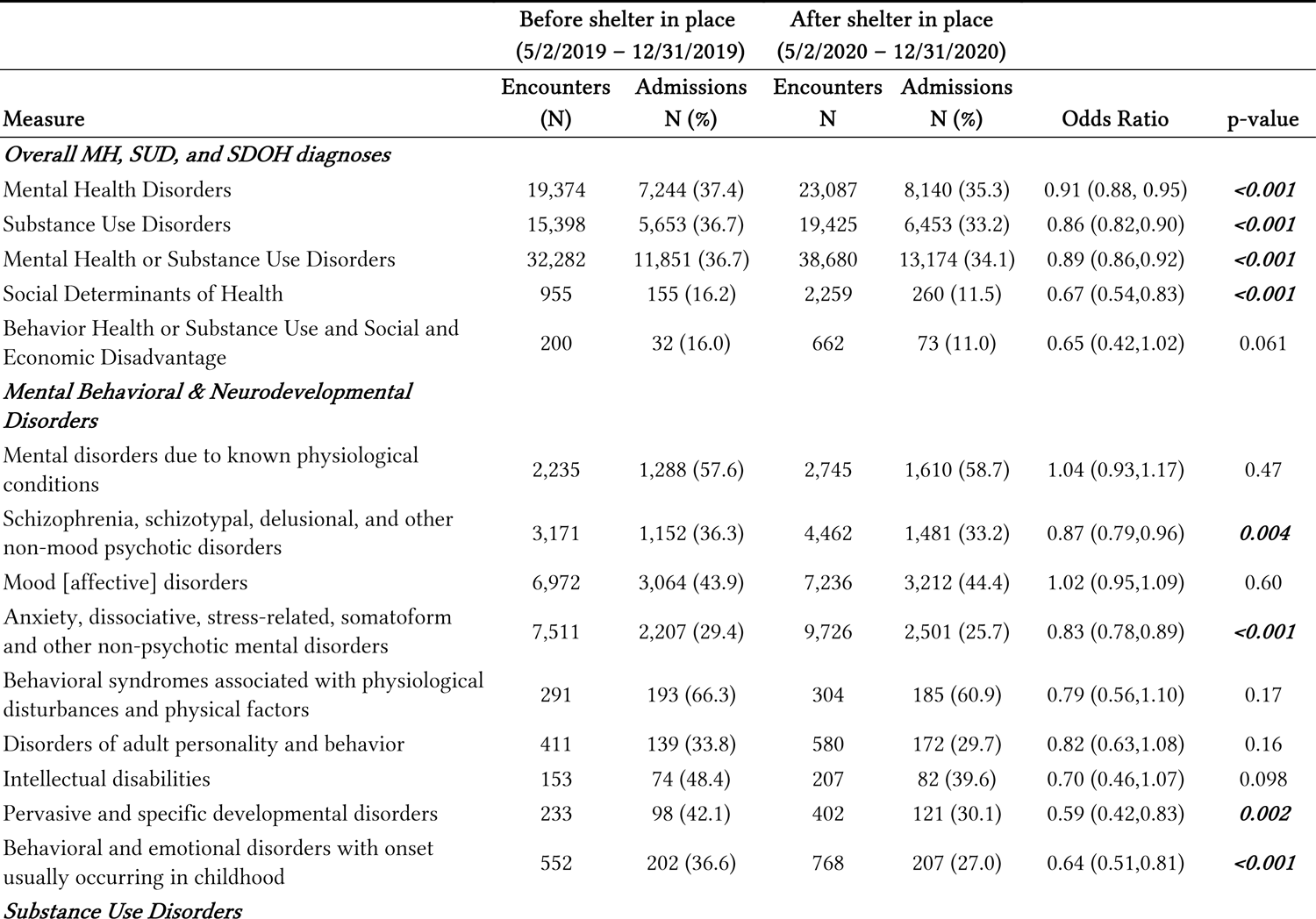
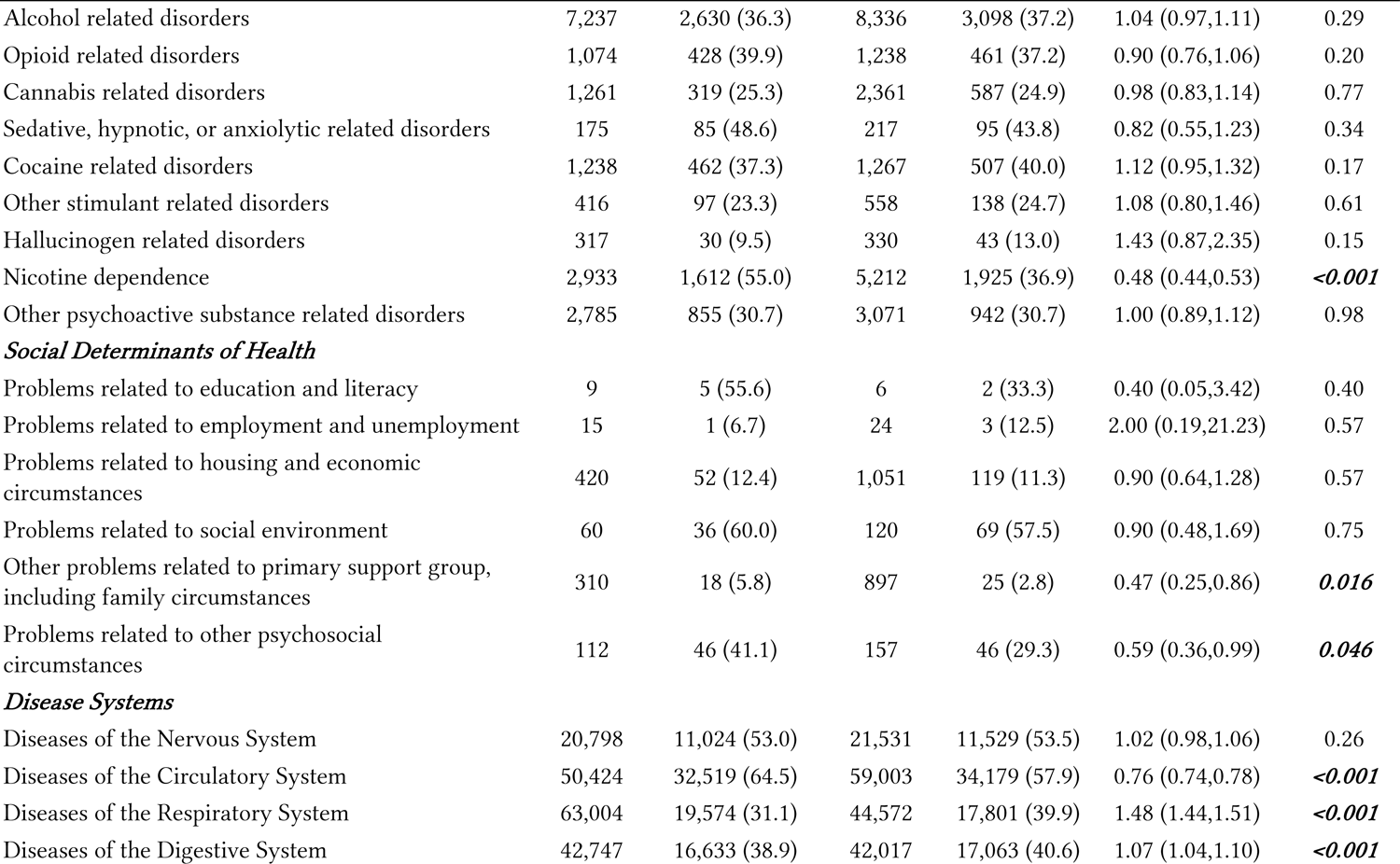
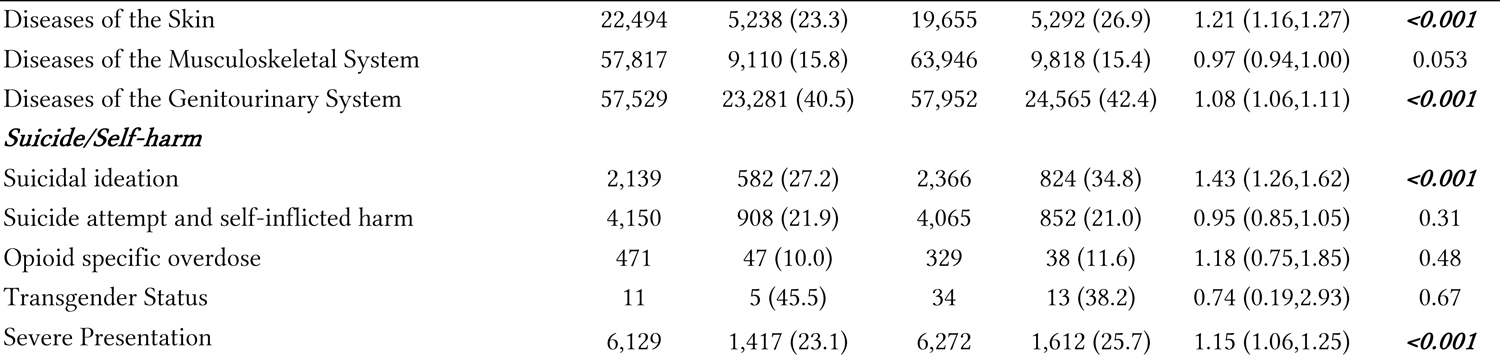
Admission rates of categories of historical diagnoses before and after shelter-in-place orders.

|  | Before shelter in place<br>(5/2/2019 – 12/31/2019) |  | After shelter in place<br>(5/2/2020 – 12/31/2020) |  |  |  |
| --- | --- | --- | --- | --- | --- | --- |
| Measure | Encounters<br>(N) | Admissions<br>N (%) | Encounters<br>N | Admissions<br>N (%) | Odds Ratio | p-value |
| *Overall MH, SUD, and SDOH diagnoses* |  |  |  |  |  |  |
| Mental Health Disorders | 19,374 | 7,244 (37.4) | 23,087 | 8,140 (35.3) | 0.91 (0.88, 0.95) | **<0.001** |
| Substance Use Disorders | 15,398 | 5,653 (36.7) | 19,425 | 6,453 (33.2) | 0.86 (0.82,0.90) | **<0.001** |
| Mental Health or Substance Use Disorders | 32,282 | 11,851 (36.7) | 38,680 | 13,174 (34.1) | 0.89 (0.86,0.92) | **<0.001** |
| Social Determinants of Health | 955 | 155 (16.2) | 2,259 | 260 (11.5) | 0.67 (0.54,0.83) | **<0.001** |
| Behavior Health or Substance Use and Social and Economic Disadvantage | 200 | 32 (16.0) | 662 | 73 (11.0) | 0.65 (0.42,1.02) | 0.061 |
| *Mental Behavioral & Neurodevelopmental Disorders* |  |  |  |  |  |  |
| Mental disorders due to known physiological conditions | 2,235 | 1,288 (57.6) | 2,745 | 1,610 (58.7) | 1.04 (0.93,1.17) | 0.47 |
| Schizophrenia, schizotypal, delusional, and other non-mood psychotic disorders | 3,171 | 1,152 (36.3) | 4,462 | 1,481 (33.2) | 0.87 (0.79,0.96) | **0.004** |
| Mood [affective] disorders | 6,972 | 3,064 (43.9) | 7,236 | 3,212 (44.4) | 1.02 (0.95,1.09) | 0.60 |
| Anxiety, dissociative, stress-related, somatoform and other non-psychotic mental disorders | 7,511 | 2,207 (29.4) | 9,726 | 2,501 (25.7) | 0.83 (0.78,0.89) | **<0.001** |
| Behavioral syndromes associated with physiological disturbances and physical factors | 291 | 193 (66.3) | 304 | 185 (60.9) | 0.79 (0.56,1.10) | 0.17 |
| Disorders of adult personality and behavior | 411 | 139 (33.8) | 580 | 172 (29.7) | 0.82 (0.63,1.08) | 0.16 |
| Intellectual disabilities | 153 | 74 (48.4) | 207 | 82 (39.6) | 0.70 (0.46,1.07) | 0.098 |
| Pervasive and specific developmental disorders | 233 | 98 (42.1) | 402 | 121 (30.1) | 0.59 (0.42,0.83) | **0.002** |
| Behavioral and emotional disorders with onset usually occurring in childhood | 552 | 202 (36.6) | 768 | 207 (27.0) | 0.64 (0.51,0.81) | **<0.001** |
| *Substance Use Disorders* |  |  |  |  |  |  |

| Measure | Before shelter in place<br>(5/2/2019 – 12/31/2019) |  | After shelter in place<br>(5/2/2020 – 12/31/2020) |  | Odds Ratio | p-value |
| --- | --- | --- | --- | --- | --- | --- |
|  | Encounters<br>(N) | Admissions<br>N (%) | Encounters<br>N | Admissions<br>N (%) |  |  |
| Alcohol related disorders | 7,237 | 2,630 (36.3) | 8,336 | 3,098 (37.2) | 1.04 (0.97,1.11) | 0.29 |
| Opioid related disorders | 1,074 | 428 (39.9) | 1,238 | 461 (37.2) | 0.90 (0.76,1.06) | 0.20 |
| Cannabis related disorders | 1,261 | 319 (25.3) | 2,361 | 587 (24.9) | 0.98 (0.83,1.14) | 0.77 |
| Sedative, hypnotic, or anxiolytic related disorders | 175 | 85 (48.6) | 217 | 95 (43.8) | 0.82 (0.55,1.23) | 0.34 |
| Cocaine related disorders | 1,238 | 462 (37.3) | 1,267 | 507 (40.0) | 1.12 (0.95,1.32) | 0.17 |
| Other stimulant related disorders | 416 | 97 (23.3) | 558 | 138 (24.7) | 1.08 (0.80,1.46) | 0.61 |
| Hallucinogen related disorders | 317 | 30 (9.5) | 330 | 43 (13.0) | 1.43 (0.87,2.35) | 0.15 |
| Nicotine dependence | 2,933 | 1,612 (55.0) | 5,212 | 1,925 (36.9) | 0.48 (0.44,0.53) | **<0.001** |
| Other psychoactive substance related disorders | 2,785 | 855 (30.7) | 3,071 | 942 (30.7) | 1.00 (0.89,1.12) | 0.98 |
| ***Social Determinants of Health*** |  |  |  |  |  |  |
| Problems related to education and literacy | 9 | 5 (55.6) | 6 | 2 (33.3) | 0.40 (0.05,3.42) | 0.40 |
| Problems related to employment and unemployment | 15 | 1 (6.7) | 24 | 3 (12.5) | 2.00 (0.19,21.23) | 0.57 |
| Problems related to housing and economic circumstances | 420 | 52 (12.4) | 1,051 | 119 (11.3) | 0.90 (0.64,1.28) | 0.57 |
| Problems related to social environment | 60 | 36 (60.0) | 120 | 69 (57.5) | 0.90 (0.48,1.69) | 0.75 |
| Other problems related to primary support group, including family circumstances | 310 | 18 (5.8) | 897 | 25 (2.8) | 0.47 (0.25,0.86) | **0.016** |
| Problems related to other psychosocial circumstances | 112 | 46 (41.1) | 157 | 46 (29.3) | 0.59 (0.36,0.99) | **0.046** |
| ***Disease Systems*** |  |  |  |  |  |  |
| Diseases of the Nervous System | 20,798 | 11,024 (53.0) | 21,531 | 11,529 (53.5) | 1.02 (0.98,1.06) | 0.26 |
| Diseases of the Circulatory System | 50,424 | 32,519 (64.5) | 59,003 | 34,179 (57.9) | 0.76 (0.74,0.78) | **<0.001** |
| Diseases of the Respiratory System | 63,004 | 19,574 (31.1) | 44,572 | 17,801 (39.9) | 1.48 (1.44,1.51) | **<0.001** |
| Diseases of the Digestive System | 42,747 | 16,633 (38.9) | 42,017 | 17,063 (40.6) | 1.07 (1.04,1.10) | **<0.001** |

| Measure | Before shelter in place<br>(5/2/2019 – 12/31/2019) |  | After shelter in place<br>(5/2/2020 – 12/31/2020) |  | Odds Ratio | p-value |
| --- | --- | --- | --- | --- | --- | --- |
|  | Encounters<br>(N) | Admissions<br>N (%) | Encounters<br>N | Admissions<br>N (%) |  |  |
| Diseases of the Skin | 22,494 | 5,238 (23.3) | 19,655 | 5,292 (26.9) | 1.21 (1.16,1.27) | **<0.001** |
| Diseases of the Musculoskeletal System | 57,817 | 9,110 (15.8) | 63,946 | 9,818 (15.4) | 0.97 (0.94,1.00) | 0.053 |
| Diseases of the Genitourinary System | 57,529 | 23,281 (40.5) | 57,952 | 24,565 (42.4) | 1.08 (1.06,1.11) | **<0.001** |
| ***Suicide/Self-harm*** |  |  |  |  |  |  |
| Suicidal ideation | 2,139 | 582 (27.2) | 2,366 | 824 (34.8) | 1.43 (1.26,1.62) | **<0.001** |
| Suicide attempt and self-inflicted harm | 4,150 | 908 (21.9) | 4,065 | 852 (21.0) | 0.95 (0.85,1.05) | 0.31 |
| Opioid specific overdose | 471 | 47 (10.0) | 329 | 38 (11.6) | 1.18 (0.75,1.85) | 0.48 |
| Transgender Status | 11 | 5 (45.5) | 34 | 13 (38.2) | 0.74 (0.19,2.93) | 0.67 |
| Severe Presentation | 6,129 | 1,417 (23.1) | 6,272 | 1,612 (25.7) | 1.15 (1.06,1.25) | **<0.001** |

## 4. Discussion

### The effect of shelter-in-place orders on vulnerable patient populations

The COVID-19 pandemic has underscored the importance of addressing social determinants of health, which are critical in shaping an individual’s health status and access to care. Our analysis of ED presentation and admission rates pre-and post-SIP orders found that patients with previously diagnosed MH or SUDs and those with a documented SDOH were more likely to present to the ED post-SIP. Prior studies that analyzed MH and substance use related ED visits demonstrate increased visit rates for these conditions as a primary diagnosis or chief complaint.^6–8^ This study extends previous work by demonstrating that SIP orders significantly impacted vulnerable patients with a history of MH, SUDs and SDOH.

Moreover, our analysis revealed that patients with co-occurring MH/SUD and SDOH had the highest increase in ED visits following SIP orders. These patients may have faced increased difficulties in accessing care and maintaining their overall health due to the closure or reduction of community-based resources^9^ and the exacerbation of existing socioeconomic inequities during the pandemic.^5^ Individuals with multiple disadvantages are at a significantly higher risk of premature mortality from avoidable causes than those with a single disadvantage^10^, highlighting the urgent need for tailored interventions that address the complex interplay of social, economic, and mental health-related factors contributing to poor health outcomes among these populations. Furthermore, patients with MH/SUDs or SDOH were less likely to be admitted to the hospital after presenting to the ED. While our study was not designed to examine the reasons behind these disparities, possible explanations include inadequate inpatient resources for patients with MH/SUD and SDOH or healthcare professional biases, as well as structural changes of healthcare resources being shunted to emergent medical care beds.

### Low documentation of social determinants of health in patients visiting the ED

Our study found that only 0.53% of ED patients had a documented Z-Code for a SDOH before and during early COVID, which is lower than other rates previously reported.^3,4,11^ Molina et al. estimated that 1.21% of ED visits have a coded SDOH; however, our paper analyzed SDOH coding at the patient level rather than the visit level, which may explain the differences observed.^12^ The usage of Z-Codes to identify patients with SDOH has repeatedly been shown to be underutilized and our results likely underestimate the number of patients with critical SDOH seeking care at EDs.^13^ Future studies could consider using natural language processing models to analyze provider notes to identify patients with SDOH that do not have a documented Z-code.

Identifying patients with SDOH is crucial because they are more likely to experience adverse health outcomes and increased ED utilization.^14,15^ Coding SDOH is currently not reimbursable, which is likely reflected in its low adoption rate; however, documentation of SDOH can substantiate patient complexity and reveal trends in healthcare utilization. For instance, if hospital systems accurately track SDOH, they can identify Z-Codes associated with higher readmission rates and allocate appropriate resources to improve patient care and outcomes. In the future, Z-Codes could play a role in determining payment rates and risk adjustment, but consistent documentation of SDOH is necessary.^16^

### Limitations

Our results should be interpreted in the context of their limitations. First, the retrospective design of our study limits our ability to draw causal conclusions. Second, our data comes from one large integrated healthcare system, which may differ from other healthcare systems or geographic regions. Nonetheless, our study includes a large and diverse patient population from different levels of care within the state. Third, our findings may be influenced by institution-specific documentation practices that could affect SDOH coding rates and limit the generalizability of our findings. Finally, our analysis relied on patients’ past medical history documentation in the EMR, which may not reflect their current diagnoses at the time of their ED visit. However, given the time constraints of the ED, we considered capturing diagnoses from past medical history the best way to represent our sample’s health status.

## Conclusion

Our study highlights the impact of the COVID-19 pandemic on vulnerable patient populations with a history of MH, SUDs, and SDOH. Our findings demonstrate that these patients were more likely to present to the ED but less likely to be admitted following SIP orders, indicating a need for tailored interventions that address the complex interplay of socioeconomic and mental health-related factors contributing to poor health outcomes among these populations. The findings highlight the need for greater standardization and consistency in documenting SDOH and addressing these factors in healthcare delivery.

## Credit author statement

**Philip Wang:** Writing – original draft preparation, methodology, conceptualization **Akhil Anand:** conceptualization, methodology, writing – review & editing, supervision **James Bena:** software, formal analysis, data curation **Shannon Morrison:** software, formal analysis, data curation **Jeremy Weleff:** conceptualization, methodology, writing – review & editing, supervision

## Abbreviations

MH: Mental health

SUD: Substance use disorder

SDOH: Social determinant of health

ED: Emergency department

SIP: Shelter in place

## Data Availability

All data produced in the present study are available upon reasonable request to the authors.

---

**Supplemental Table 1:** Counts and frequencies of patients with historical diagnoses of interest.

| Diagnosis | Total<br>(N=487,028) |
| --- | --- |
| *Overall MH, SUD, and SDOH diagnoses* |  |
| Mental Health Disorders | 49,742 (10.2) |
| Substance Use Disorders | 24,549 (5.0) |
| Social Determinants of Health, | 2,572 (0.53) |
| *Mental Behavioral & Neurodevelopmental Disorders* |  |
| Mental disorders due to known physiological conditions, | 4,134 (0.85) |
| Schizophrenia, schizotypal, delusional, and other non-mood psychotic disorders, | 4,212 (0.86) |
| Mood [effective] disorders, | 11,084 (2.3) |
| Anxiety, dissociative, stress-related, somatoform and other non-psychotic mental disorders, | 14,208 (2.9) |
| Behavioral syndromes associated with physiological disturbances and physical factors, | 541 (0.11) |
| Disorders of adult personality and behavior, | 702 (0.14) |
| Intellectual disabilities, | 250 (0.05) |
| Pervasive and specific developmental disorders, | 481 (0.10) |
| Behavioral and emotional disorders with onset usually occurring in childhood, | 1,152 (0.24) |
| *Substance Use Disorders* |  |
| Alcohol related disorders, | 10,052 (2.1) |
| Opioid related disorders, | 1,897 (0.39) |
| Cannabis related disorders, | 3,277 (0.67) |
| Sedative, hypnotic, or anxiolytic related disorders, | 379 (0.08) |
| Other stimulant related disorders, | 873 (0.18) |
| Hallucinogen related disorders, | 474 (0.10) |
| Nicotine dependence, | 7,699 (1.6) |
| Other psychoactive substance related disorders, | 4,678 (0.96) |
| *Social Determinants of Health* |  |
| Problems related to education and literacy, | 15 (0.00) |
| Problems related to employment and unemployment, | 39 (0.01) |
| Occupational exposure to risk factors, | 45 (0.01) |
| Problems related to housing and economic circumstances, | 802 (0.16) |
| Problems related to social environment, | 177 (0.04) |
| Problems related to upbringing, | 12 (0.00) |
| Other problems related to primary support group, including family circumstances | 1,157 (0.24) |
| Problems related to certain psychosocial circumstances, | 4 (0.00) |
| Problems related to other psychosocial circumstances, | 263 (0.05) |
| *Disease Systems* |  |
| Neoplasms, | 13,424 (2.8) |
| Diseases of the blood and blood-forming organs and certain disorders involving immune mechanism, | 31,867 (6.5) |
| Endocrine, nutritional and metabolic diseases, | 82,877 (17.0) |
| Diseases of the Nervous System, | 35,789 (7.3) |
| Diseases of the eye and adnexa, | 14,523 (3.0) |

| Diagnosis | Total<br>(N=487,028) |
| --- | --- |
| Diseases of the ear and mastoid process, | 14,830 (3.0) |
| Diseases of the Circulatory System, | 80,081 (16.4) |
| Diseases of the Respiratory System, | 85,219 (17.5) |
| Diseases of the Digestive System, | 68,523 (14.1) |
| Diseases of the Skin and subcutaneous tissue, | 35,984 (7.4) |
| Diseases of the musculoskeletal system and connective tissue, | 98,638 (20.3) |
| Diseases of the genitourinary system, | 84,660 (17.4) |
| Pregnancy, childbirth and the puerperium, | 676 (0.14) |
| Certain conditions originating in the perinatal period, | 1,002 (0.21) |
| Congenital malformations, deformations and chromosomal abnormalities, | 2,155 (0.44) |
| Symptoms, signs and abnormal clinical and laboratory findings, not elsewhere classified, | 267,956 (55.0) |
| Injury, poisoning and certain other consequences of external causes, | 155,944 (32.0) |
| Codes for special purposes, | 11,244 (2.3) |
| External causes of morbidity, | 21,455 (4.4) |
| Factors influencing health status and contact with health services, | 101,058 (20.7) |
| ***Suicide/Self-harm*** |  |
| Suicidal ideation, | 3,956 (0.81) |
| Suicide attempt and self-inflicted harm, | 7,879 (1.6) |
| Suicide death by underlying cause of death, | 48 (0.01) |
| Suicide deaths by method, | 48 (0.01) |
| Overdoses, | 0 (0.00) |
| Opioid specific overdose, | 732 (0.15) |
| Transgender Status, | 46 (0.01) |
| Severe Presentation, | 11,244 (2.3) |
| Behavior Health/Substance Abuse and Social and Economic Disadvantage, | 525 (0.11) |

\*Data not available for all subjects. Missing values: Other problems related to primary support group, including family circumstances = 485,871. Statistics presented as N (column %).

**Supplemental Table 2.**
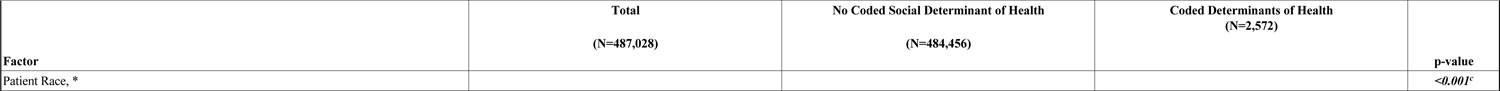
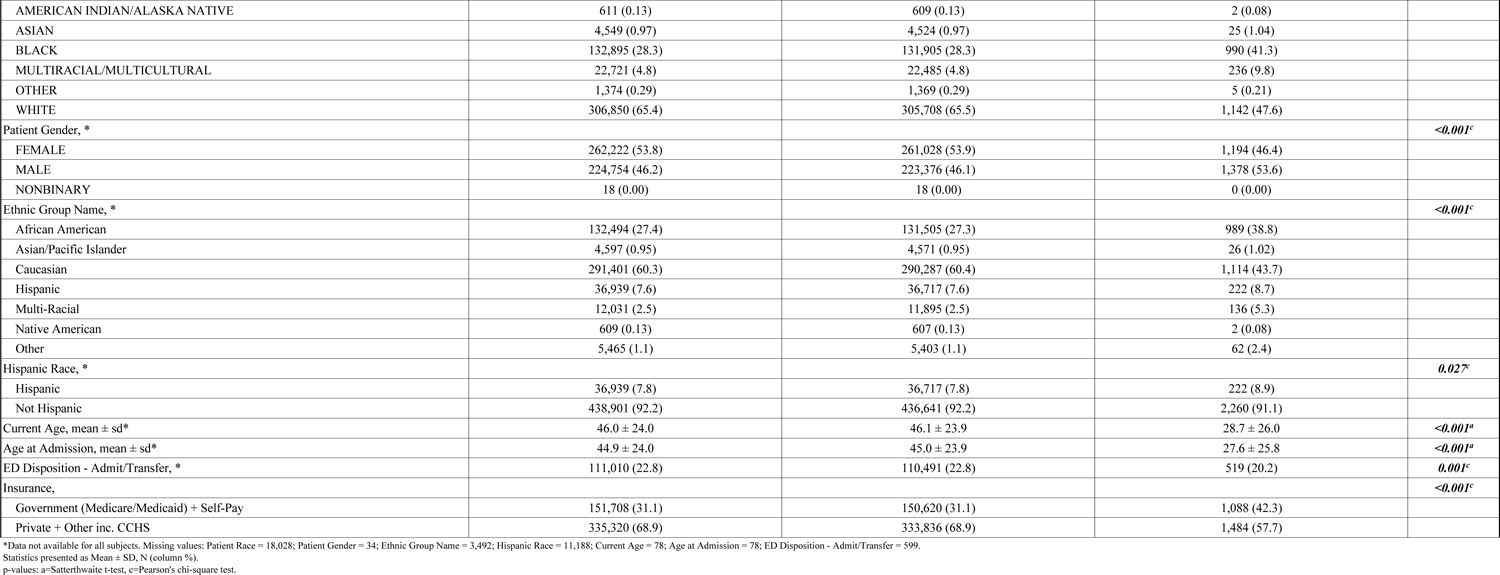
Demographics of patients with and without social determinants of health.

| Factor | Total<br>(N=487,028) | No Coded Social Determinant of Health<br>(N=484,456) | Coded Determinants of Health<br>(N=2,572) | p-value |
| --- | --- | --- | --- | --- |
| Patient Race, * |  |  |  | <0.001<sup>c</sup> |

| Factor | Total<br>(N=487,028) | No Coded Social Determinant of Health<br>(N=484,456) | Coded Determinants of Health<br>(N=2,572) | p-value |
| --- | --- | --- | --- | --- |
| AMERICAN INDIAN/ALASKA NATIVE | 611 (0.13) | 609 (0.13) | 2 (0.08) |  |
| ASIAN | 4,549 (0.97) | 4,524 (0.97) | 25 (1.04) |  |
| BLACK | 132,895 (28.3) | 131,905 (28.3) | 990 (41.3) |  |
| MULTIRACIAL/MULTICULTURAL | 22,721 (4.8) | 22,485 (4.8) | 236 (9.8) |  |
| OTHER | 1,374 (0.29) | 1,369 (0.29) | 5 (0.21) |  |
| WHITE | 306,850 (65.4) | 305,708 (65.5) | 1,142 (47.6) |  |
| Patient Gender, * |  |  |  | <0.001<sup>c</sup> |
| FEMALE | 262,222 (53.8) | 261,028 (53.9) | 1,194 (46.4) |  |
| MALE | 224,754 (46.2) | 223,376 (46.1) | 1,378 (53.6) |  |
| NONBINARY | 18 (0.00) | 18 (0.00) | 0 (0.00) |  |
| Ethnic Group Name, * |  |  |  | <0.001<sup>c</sup> |
| African American | 132,494 (27.4) | 131,505 (27.3) | 989 (38.8) |  |
| Asian/Pacific Islander | 4,597 (0.95) | 4,571 (0.95) | 26 (1.02) |  |
| Caucasian | 291,401 (60.3) | 290,287 (60.4) | 1,114 (43.7) |  |
| Hispanic | 36,939 (7.6) | 36,717 (7.6) | 222 (8.7) |  |
| Multi-Racial | 12,031 (2.5) | 11,895 (2.5) | 136 (5.3) |  |
| Native American | 609 (0.13) | 607 (0.13) | 2 (0.08) |  |
| Other | 5,465 (1.1) | 5,403 (1.1) | 62 (2.4) |  |
| Hispanic Race, * |  |  |  | 0.027<sup>c</sup> |
| Hispanic | 36,939 (7.8) | 36,717 (7.8) | 222 (8.9) |  |
| Not Hispanic | 438,901 (92.2) | 436,641 (92.2) | 2,260 (91.1) |  |
| Current Age, mean ± sd* | 46.0 ± 24.0 | 46.1 ± 23.9 | 28.7 ± 26.0 | <0.001<sup>a</sup> |
| Age at Admission, mean ± sd* | 44.9 ± 24.0 | 45.0 ± 23.9 | 27.6 ± 25.8 | <0.001<sup>a</sup> |
| ED Disposition - Admit/Transfer, * | 111,010 (22.8) | 110,491 (22.8) | 519 (20.2) | 0.001<sup>c</sup> |
| Insurance, |  |  |  | <0.001<sup>c</sup> |
| Government (Medicare/Medicaid) + Self-Pay | 151,708 (31.1) | 150,620 (31.1) | 1,088 (42.3) |  |
| Private + Other inc. CCHS | 335,320 (68.9) | 333,836 (68.9) | 1,484 (57.7) |  |

\*Data not available for all subjects. Missing values: Patient Race = 18,028; Patient Gender = 34; Ethnic Group Name = 3,492; Hispanic Race = 11,188; Current Age = 78; Age at Admission = 78; ED Disposition - Admit/Transfer = 599. Statistics presented as Mean ± SD, N (column %). p-values: a=Satterthwaite t-test, c=Pearson's chi-square test.

